# Aerobic exercise training selectively improves cortical inhibitory function after stroke

**DOI:** 10.1101/2022.11.19.22282314

**Authors:** Jacqueline A Palmer, Alicen A Whitaker, Aiden M Payne, Bria L Bartsch, Darcy S Reisman, Pierce E Boyne, Sandra A Billinger

**Affiliations:** Department of Neurology, School of Medicine, University of Kansas Medical Center, Kansas City, KS, United States of America; University of Kansas Alzheimer’s Disease Center, Fairway, KS, United States of America; Department of Physical Therapy, Rehabilitation Science, and Athletic Training, University of Kansas Medical Center, Kansas City, KS, USA; Department of Psychology, College of Arts and Sciences, Florida State University, Tallahassee, FL, USA; Department of Physical Therapy, College of Health Sciences, University of Delaware; Newark, DE; Department of Rehabilitation, Exercise and Nutrition Sciences, College of Allied Health Sciences, University of Cincinnati; Cincinnati, OH

**Keywords:** lactate, response inhibition, vigorous exercise, event-related potential, electroencephalography, executive function

## Abstract

**Background:** Aerobic exercise elicits striking effects on neuroplasticity and cognitive executive function but is poorly understood after stroke.

**Objective:** We tested the effect of 4 weeks of aerobic exercise training on inhibitory and facilitatory elements of cognitive executive function and electroencephalography (EEG) markers of cortical inhibition and facilitation. We investigated relationships between stimulus-evoked cortical responses, blood lactate levels during training, and aerobic fitness post-intervention.

**Methods:** Twelve individuals with chronic (>6mo) stroke completed an intensive aerobic exercise intervention (40-mins, 3x/week). Electroencephalography and motor response times were assessed during congruent (response facilitation) and incongruent (response inhibition) stimuli of a Flanker task. Aerobic fitness capacity was assessed as VO_2_-peak during a treadmill test pre- and post-intervention. Blood lactate was assessed acutely (<1 min) after exercise each week. Cortical inhibition (N2) and facilitation (frontal P3) were quantified as peak amplitudes and latencies of stimulus evoked EEG activity over the frontal cortical region.

**Results:** Following exercise training, the response inhibition speed increased while response facilitation remained unchanged. A relationship between earlier cortical N2 response and faster response inhibition emerged post-intervention. Individuals who produced higher lactate during exercise training achieved faster response inhibition and tended to show earlier cortical N2 responses post-intervention. There were no associations between VO_2_-peak and metrics of behavioral or neurophysiologic function.

**Conclusions:** These findings provide novel evidence for selective benefits of aerobic exercise on inhibitory control during the initial 4-week period after initiation of exercise training, and implicate a potential therapeutic effect of lactate on post-stroke cortical inhibitory function.

**Trial Registration:** ClinicalTrials.gov Identifier: NCT03760016. First posted: November 30, 2018. https://clinicaltrials.gov/ct2/show/NCT03760016

## 1. Introduction

Aerobic exercise can elicit striking effects on brain function and behavior but is poorly understood after stroke. In particular, vigorous intensity aerobic exercise has generated recent interest in the field of stroke recovery for its positive effects on neuroplasticity.^1–6^ Impaired cognitive executive function is one of the most common behavioral manifestations after stroke.^7,8^ Aerobic exercise training is widely reported as an effective approach to improve cognition, particularly for executive function processes.^9,10^ Among several domains of cognitive executive function (e.g., response inhibition, response facilitation, cognitive flexibility, planning and execution), aerobic exercise may most strongly benefit response inhibitory control that engages inhibitory cortical networks.^11–14^ The potential for aerobic exercise to improve inhibitory control is of particular interest in people after stroke who demonstrate degraded cortical inhibitory function,^15–18^ which is linked to cognitive and motor dysfunction.^15–18^ Given the well-established benefits of aerobic exercise for cardiovascular health, the effect of exercise training on persistent behavioral deficits and the dysfunctional cortical neurophysiology underlying these deficits after stroke remains elusive.

Real-time cortical processing during behavior can be probed through electroencephalographic (EEG) markers of cortical activity. EEG signatures of cortical activity are associated with clinical outcomes of post-stroke recovery^17–19^ and provide a biomarker for neural substrates that may be targeted with rehabilitation efforts.^20^ Well-characterized components of the evoked cortical activity quantified at precise timescales during behavioral task performance provide insight into distinct features of cortical facilitatory and inhibitory processes.^36–39^ Improvements in EEG markers of cortical inhibitory processing and behavioral response inhibition performance have been observed with aerobic exercise training in young, neurotypical adults.^12,28,29^ Such effects are also observed acutely after exercise in chronic stroke, in which earlier latencies and higher amplitudes of cortical activity during general attention and response inhibition tasks were observed following a single 20-minute bout of aerobic exercise.^9^ However, the chronic effect of habitual aerobic exercise training on cortical function and associated cognitive behavior have not been investigated in people post-stroke.

The intensity level of exercise training has generated recent interest across the clinical research community as an approach to optimize neuroplasticity and functional recovery after stroke.^3,30,31^ High blood lactate levels, achieved with more vigorous exercise training intensities, have been posited as a key mechanism for exercise-induced improvements in cortical function.^4,32,33^ Yet, the relationships between exercise-induced increases in blood lactate, aerobic fitness, and cortical function are unclear, particularly in the context of neurologic dysfunction after stroke. The limited understanding of these relationships presents a barrier to clinical translation, where aerobic exercise is not typically used as part of an intervention strategy to promote neuroplasticity for post-stroke recovery.^34^

In the present study, we provide an initial investigation of the effects of 4 weeks of intensive aerobic exercise training on behavioral and neurophysiologic metrics of cortical function across domains of inhibitory and facilitatory control in individuals with chronic stroke. In exploratory analyses, we probe the effects of aerobic fitness and blood lactate level produced over the course of exercise training on post-stroke behavioral and neurophysiologic cortical function. We hypothesized that 1) aerobic exercise training would elicit greater improvements in post-stroke executive function involving inhibitory compared to facilitatory control, and 2) higher blood lactate produced during exercise and greater aerobic fitness capacity post-intervention would be associated with greater cortical inhibitory function.

## 2. Methods

This study is ancillary to a primary multi-site randomized control trial (RCT).^31^ A detailed trial protocol has been described in Miller et al. (2021)^35^ and primary endpoint analyses for the full participant cohort are described in Boyne et al., 2022.^31^ Briefly, participants (40-80 years old) sustained a single stroke 6mo-5yrs prior to consent, could walk at least 10 meters with no physical assistance, were of stable cardiovascular condition, and possessed no other neurologic diagnoses or significant musculoskeletal pain with exercise.^31^ The experimental protocols for the primary and ancillary study protocols were approved by the University of Kansas Medical Center Institutional Review Board and all participants provided written, informed consent (UC IRB 2017-5325; KUMC IRB MOD00023271; STUDY00143669).

### 2.1. Participants

A subgroup of 16 eligible and interested participants from the University of Kansas Medical Center participated in the present ancillary study. In addition to clinical and aerobic fitness assessments associated with the primary study, these participants were asked to complete cognitive assessments with concurrent electroencephalography (EEG) at baseline and 2-7 days after the final exercise training session within the 4-week intervention.

### 2.2. Exercise intervention

The exercise intervention methods are thoroughly detailed in the protocol and primary outcomes papers for the present study.^31,35^ Briefly, following baseline assessments participants were randomized to exercise training consisting of either high-intensity interval or moderate-intensity continuous walking exercise intervention arms. The aim of this ancillary study was to investigate behavioral and neurophysiologic metrics of cortical function with exercise training, thus participants from both intensity groups were pooled together. This grouped-analytic approach provided a greater range of data points from which the effects tested in the present study could be elucidated, and has been used previously in post-stroke investigations of changes in physiologic metrics with exercise training.^36^ As published previously, all participants in each arm completed 40-minutes total of mixed over-ground and treadmill walking, which was either continuous and targeting ∼50% age predicted heart rate reserve in the moderate-intensity group or bursts of 30 s of walking at the maximum safe speed separated by ∼30-60s of resting in the high-intensity interval group, 3x/week.^31,35^ After 4 weeks of training (12 sessions) post-intervention assessments were performed within 2-7 days by the same blinded experimenter.

### 2.3. Cardiovascular fitness and clinical walking assessment

Participants completed a treadmill graded exercise test (GXT) during which oxygen consumption (VO_2_) was continuously measured.^31^ Aerobic fitness was quantified as the peak VO_2_ achieved during the GXT assessment. Self-selected and maximal walking speed also were assessed using the 10-meter walk. Functional walking capacity, the primary outcome of the parent study,^31^ was assessed using a 6-minute walk test. Assessments were performed at baseline and post-intervention after 4 weeks of exercise training.

### 2.4. Blood lactate measures of exercise intensity

Blood lactate concentration was rapidly assessed (<1 minute) after treadmill exercise cessation using a finger stick method and a point-of-care blood lactate analyzer (Lactate Plus, Nova Biomedical, Waltham, MA, USA) at the mid-week training session each week. The mean blood lactate during exercise training over the 4 weeks for each individual was used for analyses.

### 2.5. Eriksen Flanker task

Participants completed a modified arrow Eriksen Flanker task,^37^ during which cognitive executive function behavior and concurrent EEG recordings of response facilitation and inhibition were measured. During the task, participants were seated comfortably in a chair with both hands resting on a table in front of them and a computer mouse control positioned under the index and middle fingers of the nonparetic hand. During each trial, a computer monitor displayed five arrowheads at eye-level. Participants were instructed to press the left or right mouse button corresponding to the direction of the central arrowhead. There were two “congruent” stimuli (“<<<<<“ and “>>>>>“) and two “incongruent” stimuli (“<<><<“ and “>><>>“). On each trial, the stimulus was presented for 200ms, and the inter-stimulus interval was jittered randomly between 3000 to 3200ms. Participants were instructed to complete the task as quickly and accurately as possible. Each participant first completed a practice block of 12 trials consisting of randomized delivery of 3 trials of each left and right congruent and incongruent stimuli to ensure that they understood the task. Following the practice block, 200 stimuli were presented randomly, with 50 trials for each left/right directions and congruent/incongruent conditions (total 200 trials).

#### 2.5.1. Behavioral performance during the Flanker task

Performance metrics of accuracy and response time were assessed separately for congruent and incongruent stimuli. Accuracy was quantified as the percentage of correct responses out of total responses. Response latencies were assessed in the congruent condition, in which participants facilitated a rapid response clearly indicated by the stimulus, and in the incongruent condition, in which participants needed to rapidly inhibit the prepotent, incorrect response that is activated by the conflicting stimulus information. Trials with incorrect responses, indicating failure to inhibit the incorrect prepotent responses, were excluded from measurement of response latencies.^9^ Response latencies were quantified as the mean time between visual stimulus onset and accurate motor response across all correct trials for each condition. The response time cost of inhibiting the prepotent incorrect response to the incongruent stimulus (response inhibition) was quantified within individuals as the relative increase in response time latency during the incongruent condition as: response time (incongruent)/response time (congruent).

#### 2.5.2. Electroencephalography (EEG) data collection and analyses

During the Flanker task, EEG recordings of cortical activity were acquired using 256-channel HydroCel Geodesic Sensor Net (Electrical Geodesics, Inc.) sampled at 1000 Hz and stored for offline analysis. All data processing and analyses were performed using freely available functions from the EEGlab toolbox^38^ and custom MATLAB software. Time-locked continuous data were imported into EEGlab and filtered with a high-pass cutoff of 0.1Hz and a low-pass cutoff of 30Hz. Next, the events with trigger labels for correct trials (i.e., accurate responses) were selected and epoched at -1 to 2 s relative to the time of each visual stimulus at t=0. The EEG data were re-referenced to the average activity recorded from the left and right mastoid electrodes. Eye movement and blink artifacts were identified and corrected using the automatic method developed by Gratton, Coles, and Donchin.^39^ Trials with other artifacts were automatically detected and rejected if the range or variance of the voltage within a trial was > 2 standard deviations above the average from the same individual and channel. The data were then averaged across all trials within each condition and the average voltage between 100-400ms before stimulus onset was subtracted. Average activity at fronto-central and midline vertex sites (i.e., AFz, Fz, FCz, Cz, CPz, and Pz) and the absolute magnitudes of the peak negative (200-300ms) and positive (300-1000ms) components of the average waveform were determined for each location. Consistent with previous research demonstrating the maximal and most reliable evoked cortical responses at the fronto-central scalp region,^40^ we found the maximal and most robust evoked cortical responses for both the negative (N2) and positive (P3) components occurred at the Fz location, which was selected for further analyses. Cortical inhibitory and facilitatory processing were quantified as the latencies and amplitudes of the evoked cortical N2^21^ and P3^9,22^ components. The N2 was identified as the peak negative evoked cortical activity between 200-300ms in incongruent trials and visually inspected to confirm accuracy. Two participants had N2 responses that clearly occurred later, outside this window; for these participants, the N2 time window was shifted to 250-350ms to capture the negative peak. The P3 was identified as the peak positive evoked cortical activity between 300-1000ms in congruent trials;^19^ visual inspection confirmed reliable detection of the P3 response for each participant within this time window, consistent with previous reports in individuals post-stroke.^9^ The mean of the peak amplitudes and latencies of the N2 and P3 components across like conditions were measured at baseline and post-intervention for each participant.

Response inhibition measured during the incongruent stimuli condition is reflected in the cortical N2 component of the evoked EEG waveform; the incongruent response time and cortical N2 response latency have excellent internal consistency and moderate-to-excellent test-retest reliability during this task.^41^ Response facilitation during the congruent stimuli condition is reflected as the cortical P3 response; these metrics have excellent internal consistency and acceptable-to-good test-retest reliability.^42^

### 2.6. Statistical analyses

We tested for normality and heterogeneity of variance of all data used for analyses using Kolmogorov-Smirnov and Levene’s tests, respectively. Repeated one-way analysis-of-variance (ANOVA) tests were used to test for a change in behavioral response accuracy and speed as well as cortical response latencies and amplitudes of the N2 and P3 between baseline and post-intervention timepoints. Pearson product-moment correlation coefficients were used to test for associations between inhibitory behavioral response time cost and the N2 response latencies and amplitudes during the incongruent condition, and between behavioral response times and P3 response latencies and amplitudes during the congruent condition at baseline and at the 4-week post-intervention timepoint. In exploratory analyses, we used Pearson product-moment correlation coefficients to test the relationship between blood lactate during exercise training and peak-VO2 versus behavioral (response time, inhibitory response time cost) and cortical activity (N2, P3) components post-intervention. All analyses were performed using SPSS version 27 with an a priori level of significance set to 0.05.

## 3. Results

Of the total 16 participants who enrolled, two participants withdrew from the primary RCT within the first 4 weeks and did not complete the intervention. (**Supplementary Figure S.1**). Incomplete datasets were excluded from the present analyses. Two additional participants were unavailable for cognitive/neurophysiologic testing at the 4-week post-intervention assessment. Complete datasets were available for twelve participants (age 61±11yrs, range 47-77yrs) who completed 4 weeks of exercise training and all baseline and post-intervention assessments (**Table 1**). No serious study-related adverse events occurred during this ancillary study.

**Table 1.** Participant characteristics. PSD = Post-stroke duration.

| Participant ID | Sex | PSD (months) | Lesion Type | Lesion Side | Lesion Location |
| --- | --- | --- | --- | --- | --- |
| S01 | Female | 12.0 | Ischemic | Right | Cerebellar |
| S02 | Female | 30.0 | Ischemic | Right | Pontine |
| S03 | Male | 12.0 | Hemorrhagic | Right | Intraparenchymal |
| S04 | Male | 57.6 | Ischemic | Right | Posterior frontal periventricular white matter, putamen, and external capsule |
| S05 | Male | 30.0 | Hemorrhagic | Left | Middle cerebral artery with subarachnoid hemorrhagic conversion |
| S06 | Male | 12.0 | Ischemic | Right | Lacunar, caudate, posterior internal capsule, and posterior putamen |
| S07 | Male | 50.4 | Hemorrhagic | Left | Basal ganglia |
| S08 | Female | 20.4 | Ischemic | Right | Basal ganglia, lacunar |
| S09 | Male | 50.4 | Ischemic | Left | Middle cerebral artery |
| S10 | Male | 16.8 | Ischemic | Left | Middle cerebral artery |
| S11 | Female | 12.0 | Hemorrhagic | Right | Thalamic, intraparenchymal |
| S12 | Male | 16.8 | Hemorrhagic | Left | Basal ganglia |
| Mean (SD) | M = 8 | 26.7 (17.1) | I = 7 | L = 5 |  |

### 3.1. Aerobic fitness and clinical walking response to exercise intervention

Reporting of primary and secondary clinical outcomes for the full participant cohort are detailed in Boyne et al (2022).^31^ The subset of participants in the present ancillary study produced a mean blood lactate level of 2.42±1.35 mmol/L [Range: 1.20–5.55 mmol/L] during the first 4-week block of exercise training. Comfortable and maximal walking speed increased 4 weeks post-intervention (self-selected: baseline0.75±0.26 m/s, post 0.80±0.27 m/s, *p*=0.037; fastest: baseline 1.02±0.36 m/s, post 1.20±0.57 m/s, *p*=0.041). Functional walking capacity on the 6-minute walk test showed a mean increase that did not reach our adopted level of significance (baseline 276.7±104.6 m, post 311.2±136.1 m, *p*=0.122). Aerobic fitness capacity, measured as VO_2_-peak during the GXT showed no change after 4 weeks post-intervention (**Table 2**, baseline 15.7±3.9 mL/kg/min, post 16.2±4.3 mL/kg/min, *p*=0.288).

**Table 2.** Participant clinical and fitness outcomes in response to the intervention.VO_2_-peak: peak oxygen consumption; 6MWT: 6-minute walk test; 10MWT: 10-meter walk test.

| Table 2. |  |  |  |  |  |  |  |
| --- | --- | --- | --- | --- | --- | --- | --- |
|  | VO<sub>2</sub>-peak<br>(ml/kg/min) |  | 6MWT<br>(meters) |  | Fastest 10MWT speed<br>(meters/sec) |  | Blood<br>Lactate<br>(mmol/L) |
| Participant ID | Baseline | Post | Baseline | Post | Baseline | Post | Average |
| S01 | 18.6 | 17.4 | 400 | 271.5 | 1.47 | 2.03 | 1.43 |
| S02 | 14.5 | 13.9 | 350.0 | 399.1 | 1.20 | 1.32 | 2.23 |
| S03 | 8.8 | 10.1 | 186.9 | 224 | 0.69 | 0.73 | 1.53 |
| S04 | 15.7 | 14.5 | 309.5 | 331.0 | 1.08 | 1.14 | 2.35 |
| S05 | 19.8 | 20.9 | 450.0 | 556.5 | 1.51 | 2.04 | 5.55 |
| S06 | 13.3 | 15.0 | 168.8 | 152 | 0.99 | 0.81 | 4.45 |
| S07 | 16.7 | 21.1 | 352.0 | 424.5 | 1.40 | 1.85 | 2.00 |
| S08 | 9.0 | 9.6 | 98.8 | 88.0 | 0.32 | 0.30 | 1.25 |
| S09 | 17.0 | 17.3 | 250.0 | 288.1 | 0.86 | 0.72 | 2.65 |
| S10 | 21.6 | 24.1 | 314.1 | 421.5 | 1.16 | 1.37 | 2.95 |
| S11 | 16.2 | 15.8 | 259.8 | 400.0 | 0.94 | 1.41 | 1.50 |
| S12 | 16.6 | 15.0 | 180.2 | 178.4 | 0.65 | 0.66 | 1.20 |
| Mean (SD) | 15.7 (3.8) | 16.2 (4.3) | 276.7 (104.6) | 311.2 (136.1) | 1.02 (0.36) | 1.20 (0.57) | 2.42 (1.35) |

### 3.2. Effect of aerobic exercise training on cognitive behavior and cortical processing

After completing 4 weeks of aerobic exercise training, participants achieved a lower response time cost of inhibiting prepotent responses during incongruent stimuli (response inhibition) post-intervention (**Figure 1A**, *p*= 0.037), with no significant change in accuracy (**Figure 1B**, *p*= 0.088). Response times and accuracy during congruent stimuli (response facilitation) did not change from baseline to post-intervention (response time, *p*=0.288; accuracy, *p*=0.389). Although no group-level changes in cortical N2 or P3 responses were observed from baseline to post-intervention (latency N2, *p*=0.193; P3, *p*= 0.497; amplitude N2, *p*=0.344; P3, *p*=0.105), a positive association between the N2 latency and inhibitory response time cost emerged post-intervention (**Figure 2A&C**, *r*=0.745, *p*=0.005) that was not present at baseline (**Figure 2B**, *r*=0.088, *p*=0.780). There were no associations between amplitude of the N2 peak during the incongruent condition, or the amplitude or latency of the P3 peak during the congruent condition and concurrent behavioral response times at any time point.

**Figure 1.**
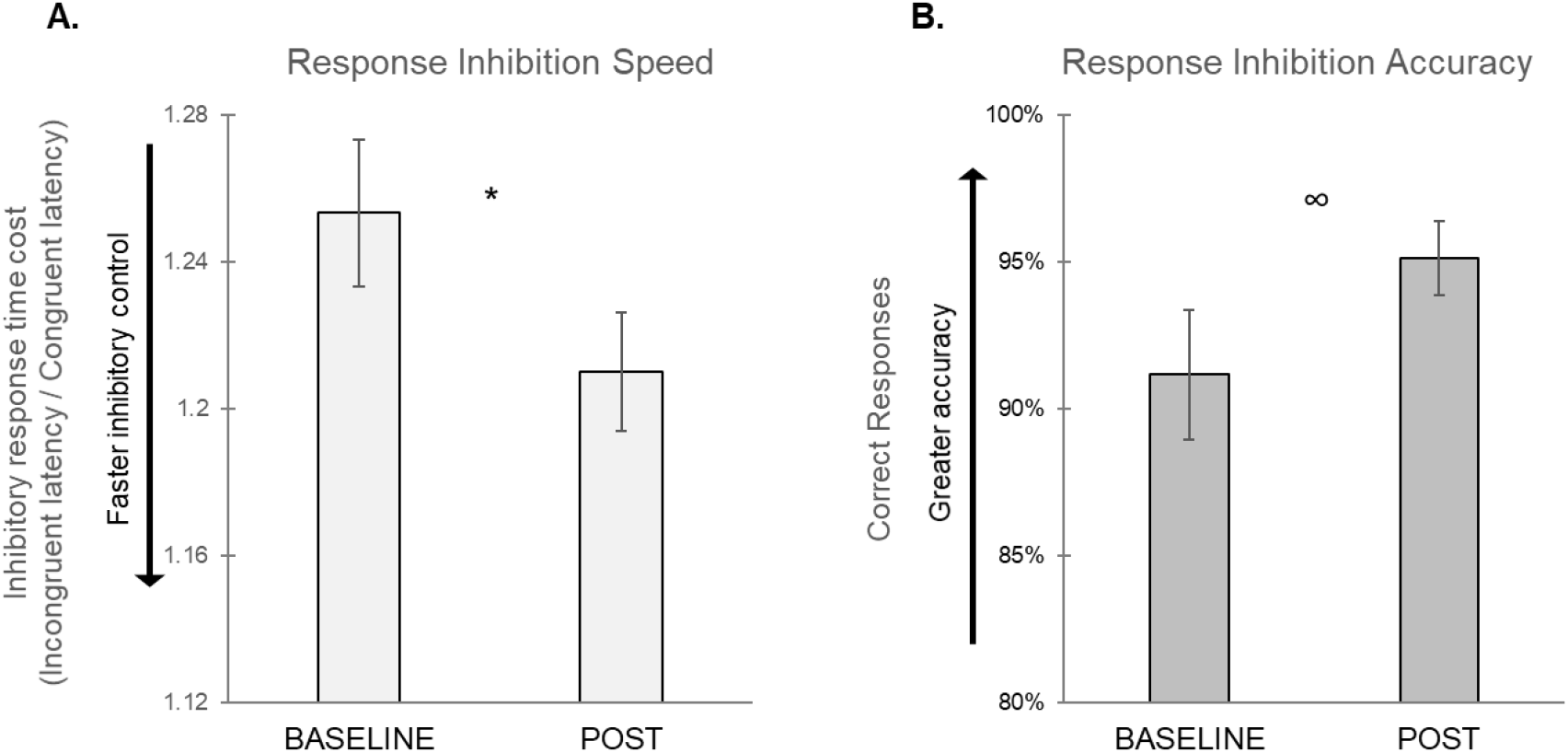
Inhibitory response time cost (A) and accuracy (B) during the incongruent stimuli condition at baseline and post-intervention (POST). The inhibitory response time cost decreased (*p=.037) across the group while response accuracy showed no significant change (∞ p=.088) post-intervention.

**Figure 2.**
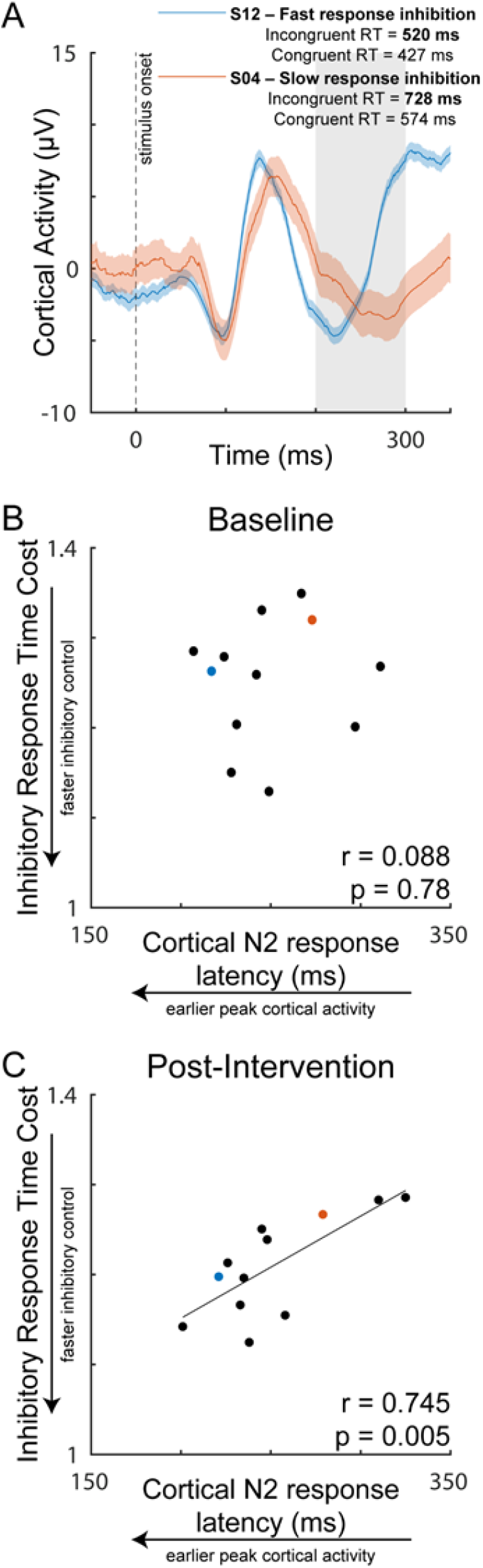
Cortical N2 response as a function of inhibitory response time cost during the incongruent stimuli condition post-intervention. Representative individuals with slow and fast response inhibition following 4 weeks of exercise training **(A)**. While no relationship was present at baseline (**B**), a positive relationship between cortical N2 response latency and inhibitory response time cost (reaction time of incongruent/congruent condition) emerged post-intervention, in which individuals with earlier peak cortical N2 responses had faster inhibitory control (**C**). The dashed line represents the time of visual stimulus onset (t=0). The shaded region represents the time window used for detection of the peak N2 response. RT = response time.

### 3.3. Effects of exercise intensity on cognitive behavior and cortical processing

Individuals who produced higher blood lactate levels during exercise training had lower inhibitory response time costs post-intervention (**Figure 3A&B**, *r*=-0.685, *p*=0.014). Similarly, we observed a pattern in which individuals with higher blood lactate had earlier cortical N2 response latencies during the incongruent condition post-intervention, though this association failed to meet our adopted level of significance (**Figure 3C**, *r*=-0.480, *p*=0.110). There were no associations between lactate and congruent response time (*r*=0.463, *p*=0.130) or P3 response latency (*r*=0.022, *p*=0.945) or amplitude (*r*=0.357, *p*=0.255).

**Figure 3.**
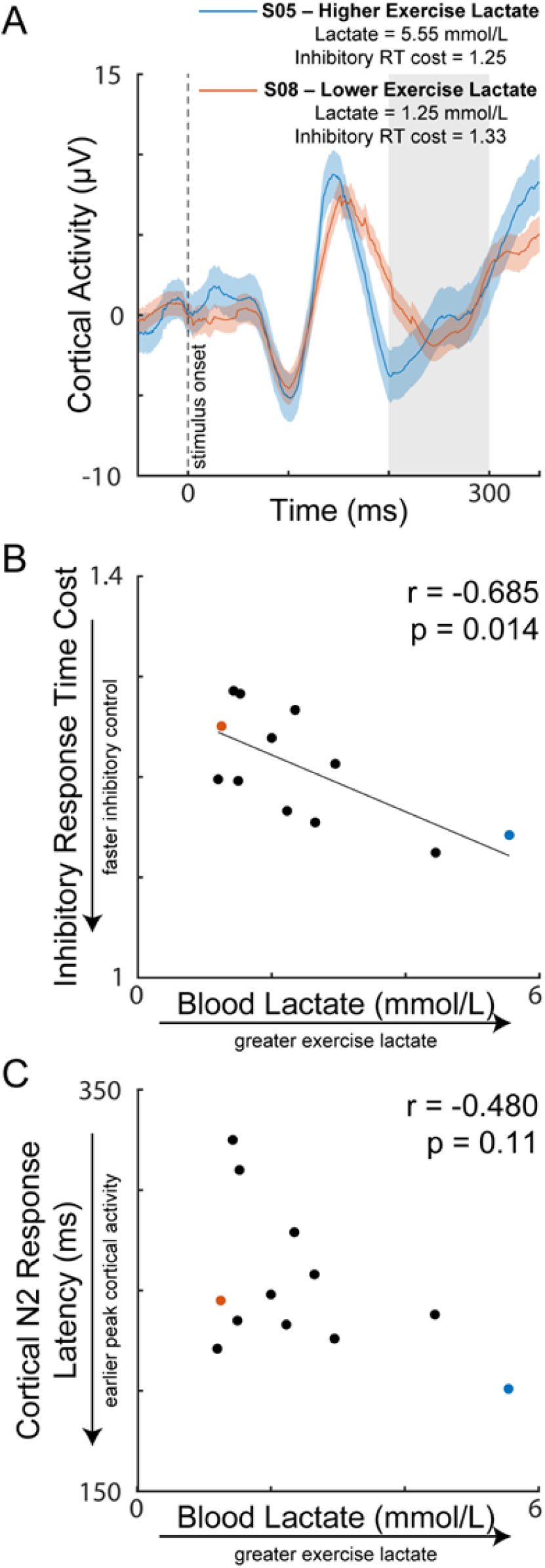
Cortical N2 response post-intervention as a function of blood lactate produced during exercise training over the course of the 4-week intervention. Representative cortical responses in individuals with higher and lower exercise blood lactate levels (**A**). There was a negative relationship between blood lactate and inhibitory response time cost (reaction time of incongruent/congruent condition), in which individuals who produced greater exercise lactate achieved faster inhibitory control performance (**B**). Individuals who produced greater exercise lactate also showed a pattern of earlier peak cortical N2 activity post-intervention, but this effect failed to meet our adopted level of significance (**C**).

### 3.4. Effects of aerobic fitness capacity on cognitive behavior and cortical processing

We observed no associations between VO_2_-peak and any domain of executive function performance (congruent response time, *p*=0.788; inhibitory response time cost, *p*=0.392) or latency or amplitude of the cortical N2 (latency, *p*=0.245; amplitude, *p*=0.177) or P3 (latency, *p*=0.418; amplitude, *p*=0.745) post-intervention.

## 4. Discussion

Findings of the present study provide initial evidence that intensive aerobic exercise training may preferentially benefit post-stroke cortical inhibitory function and strengthen neurophysiologic brain-behavior relationships in chronic stroke. Our preliminary results show that individuals who produced higher blood lactate during exercise training had faster response inhibition post-intervention, supporting further mechanistic investigation of lactate as a mediator for inhibitory cortical plasticity in individuals post-stroke. These effects occurred after only 4 weeks of exercise training and were independent of aerobic fitness capacity, suggesting that neuromotor rather than cardiovascular changes with aerobic exercise training drive improvements in post-stroke inhibitory control.

### 4.1 Aerobic exercise training selectively improves response inhibition after stroke

Our results reveal measurable improvements in post-stroke executive function after aerobic exercise training, an effect specific to behavior involving inhibitory control. We found a reduction in the response time cost of inhibiting the prepotent incorrect response to incongruent stimuli post-intervention (**Figure 1A**), without changes in general response facilitation. Interestingly, this selective effect on inhibition was not reported acutely after a single bout of aerobic exercise in stroke, in which both cortical facilitatory and inhibitory processes were modulated.^3,9^ Thus, the present findings build upon previous studies of acute, exercise-induced neuroplasticity post-stroke, suggesting a shift from a generalized to a more specific effect on inhibition over time with repeated exposure to aerobic exercise training. The present inhibitory specificity of habitual exercise training in stroke is consistent with previous reports in neurotypical populations of young ^11,12,43^ and older adults.^44–46^ For example, older adults with and without Parkinson’s disease demonstrated selectively improved cognitive inhibitory control following a 3-month aerobic exercise intervention.^44^ Notably, selective behavioral improvements in inhibitory control following aerobic exercise have been reported even in the absence of a group-level changes in the latency or amplitude of the cortical N2 response in neurologically-intact individuals,^12^ similar to the present findings in individuals post-stroke. These findings support the notion that facilitatory and inhibitory elements of cognitive executive function differentially respond to aerobic exercise training and implicates the value of assessing distinct elements of cortical facilitation and inhibition in response to aerobic exercise training.

The relationship between neurophysiologic markers of cortical processing and response inhibition behavior were strengthened after exercise training, illuminating potential mechanisms underpinning improved inhibitory control with exercise after stroke. While no relationship was present at baseline (**Figure 2B**), we found a positive relationship between cortical and behavioral indices of inhibitory functional after 4 weeks of intensive aerobic exercise training (**Figure 2B**). Specifically, individuals with earlier cortical N2 response latencies post-intervention also achieved lower inhibitory response time costs (**Figure 2A**), suggesting a strengthened link between cortical processing and resulting behavioral aspects of response inhibition. The emergence of this brain-behavior correlate with aerobic exercise training offers a mechanistic explanation for the improved cost of response inhibition post-intervention (**Figure 1A**). The strength of neurophysiologic brain-behavior relationships is behaviorally relevant in the context of aging and neuropathology, where there is a weakened association between EEG markers of cortical activity and concurrent behavior.^47–49^ The association between cortical activity and response inhibition behavior can be improved with intervention, and appears to reflect reallocation or strengthening of cortical resources to inhibitory control processes involving frontal-motor regions to detect and inhibit response conflict.^47,50,51^ Aerobic exercise training may trigger a similar shift in the neural strategy that individuals post-stroke utilize for inhibitory control, increasing the proficiency of frontal-motor cortical inhibitory processes and improving cortical reactivity mechanisms that are commonly impaired after stroke.^17,18^ After stroke, higher levels of physical activity have been linked to greater myelination within the brain,^52^ consistent with the temporal cortical N2 component (i.e. N2 latency) association with behavior following exercise training in the present study (**Figure 3**). Future studies employing multimodal neuroimaging approaches (e.g., functional magnetic resonance imaging– EEG) could provide spatial insight into the neural substrates underpinning behaviorally-salient markers of cortical inhibition and may inform the development of targeted synergistic neurorehabilitation approaches (e.g. noninvasive brain stimulation)^53^ with aerobic exercise training.

### 4.2. Lactate may facilitate benefits of aerobic exercise on cortical inhibitory function

Our findings show preliminary evidence that lactate may play a role in exercise-induced improvements in cortical inhibitory function after stroke. In the present study, individuals who produced a higher mean lactate level during 4 weeks of exercise training had lower inhibitory response time cost and tended to evoke earlier cortical N2 responses post-intervention (**Figure 3**). Notably, only two individuals reached a mean exercise lactate level of >4.0 mmol/L, the typical threshold for transition from aerobic to more anaerobic metabolism;^54^ these individuals strongly influenced the effect on lactate on behavioral and neurophysiologic markers of cortical inhibitory function (**Figure 3B&C**)and may support the salience of this threshold for the therapeutic effects of exercise lactate on post-stroke neuroplasticity. A mechanistic link between lactate and cortical inhibition, largely mediated through gamma-aminobutyric acid (GABA),^5,6^ is supported by the preferential activation of GABAergic networks after exercise by the peripheral release of lactate that crosses the blood brain barrier.^5,6^ The ability to augment GABAergic plasticity is of particular interest after stroke, where GABA modulation was associated with improved motor function following rehabilitation (e.g., constraint-induced movement therapy).^55^ The present preliminary findings warrant future studies to test if exercise lactate levels >4.0mmol/L trigger more robust effects on cortical inhibitory function, and whether lactate may target complementary GABAergic neuroplasticity induced by skilled motor practice, potentially augmenting post-stroke rehabilitation outcomes.

Heightened neuro-availability of GABA following exercise is greater after vigorous-intensity exercise producing higher blood lactate levels,^5,6^ and specifically primes neuroplasticity involving inhibitory control.^56–58^ In neurotypical, young adults, vigorous-intensity exercise elicited more robust modulation of frontal-region cortical activity and associated increases in behavioral response inhibition compared to lower exercise intensity.^4^ This targeted GABAergic effect of vigorous intensity exercise may explain the present selective effects of exercise on post-stroke inhibitory control (**Figures 1 & 3**), and raises the possibility that interventions aimed at producing sufficiently high lactate levels (e.g.,>4.0mmol/L) could magnify the observed inhibitory behavioral effects and potentially detect a group-level effect for cortical N2 response latency change with exercise training. While the pattern between higher lactate and earlier cortical N2 response did not reach statistical significance (**Figure 3C**), future studies investigating more specific indices of GABAergic network activity (e.g., beta power)^16^ and connectivity (e.g., beta coherence)^17^ may be better positioned to detect such associations. These preliminary findings provide an exciting first-step for future investigation of effects of exercise dosing and the production of peripheral metabolites on post-stroke cortical plasticity and inhibitory control.

### 4.3. Initial effects of aerobic exercise on cortical inhibitory function occur independently of aerobic fitness changes post-stroke

The present findings reveal that improvements in inhibitory control occur after only 4 weeks of habitual aerobic exercise in individuals with chronic stroke (**Figure 1**). These initial post-stroke behavioral changes and associated markers of cortical inhibition (**Figure 2**) appear to be independent of exercise-induced changes in aerobic fitness capacity, as there were no relationships between VO_2_-peak and cortical or behavioral metrics and no detectible group-level increase in VO_2_-peak post-intervention. This was an unexpected finding given the well-established link between physical fitness and cognitive function,^59^ and the older (age 61±11 years), deconditioned baseline state (VO_2_-peak=15.3±3.9 mL/kg/min) of our post-stroke participant cohort (**Tables 1& 2**). While the small sample side and short assessment timeframe may have been underpowered and insufficient to detect changes in VO_2_-peak, the observed relationships with blood lactate suggest that cortical inhibitory function can improve with exposure to sufficient production of blood lactate during the initial period of exercise training before overall changes in aerobic fitness are achieved. Our findings show that early improvements in executive inhibitory control induced by exercise training occur on a similar timescale to significant increases in short-distance gait speed (**Table 2**), that appear to be driven by neuromotor rather than cardiopulmonary adaptations.^31^ Our results imply that individuals across a spectrum of aerobic fitness levels after stroke may achieve early neuromotor benefits from intensive aerobic exercise training for speed-dependent domains of specific aspects of cognitive function and ambulatory capacity. Future studies in a larger cohort of individuals post-stroke may test for common neurophysiologic underpinnings of these initial exercise-induced increases in the speed of cognitive processing and whole-body motor behaviors such as walking.

### 4.4. Strengths and limitations

This study leveraged a multisite, RCT involving aerobic exercise training^31^ to provide novel, mechanistic insight into the effects of aerobic exercise on cortical processing and associated cognitive behavior. The exercise training employed by the present study was more intensive than that of conventional post-stroke rehabilitation and did not include skilled motor practice, which positioned us well to investigate aerobic exercise-induced effects on neurophysiologic and behavioral indices of cortical function.

Given the small sample size and preliminary nature of this study, our analyses utilized a pointed, hypotheses-driven approach that was motivated by our previous work and specific interest in post-stroke cortical inhibitory function and effects of exercise and training intensity. Thus, it is possible that we were not adequately powered to detect exercise-induced changes in cortical facilitatory function with smaller effect sizes. This small-scale ancillary study lacked a control condition (i.e. non-exercise), limiting interpretation for the causality of the exercise intervention on the results. No long-term follow-up testing occurred to determine whether improved inhibitory response time cost was further enhanced with additional training as part of the primary study RCT (e.g. 8-weeks, 12-weeks of training)^31^ or whether there were sustained effects after completing the intervention. Thus, results should be interpreted cautiously and a larger RCT with controls, additional assessments, and follow-up retention testing would be needed to confirm and expand these findings. Larger sample sizes would also provide the opportunity to test the effect of habitual aerobic exercise across a more expansive battery of cognitive domains and sophisticated analyses of cortical function (e.g., spectral and component features of EEG activity).

The nonparetic hand was used to perform executive function assessments in the present study, allowing us to control for severity of paretic upper extremity hemiparesis. We also used an established, standardized approach involving a channel-based, centralized, frontal region location analyses of cortical responses evoked during the Flanker task.^9,42^ As such, future studies are needed to test whether there is a hemispheric lateralization effect for exercise-induced improvements in post-stroke cortical inhibitory mechanisms.

We cannot rule out the potential effects of brain derived neurotrophic factor (BDNF) and other peripherally-induced neurotrophic factors that increase collaterally with lactate during exercise,^3^ on the present results. For example, Boyne et al. (2019) found that acute circulating levels of BDNF after aerobic exercise were greater after high-intensity exercise and associated with greater changes in post-stroke intracortical inhibition.^3^ However, Charalambous et al (2018) reported a mean blood lactate of 6.1 mmol/L acutely after exercise in the absence of change in circulating BDNF in individuals post-stroke, providing evidence that exercise-induced increases in lactate can occur independently of BDNF in post-stroke populations.^60^ Future studies investigating a spectrum of peripheral and muscle-induced factors and neurotrophins will enhance our mechanistic understanding of the acute and long-term effects of exercise on post-stroke neuroplasticity.

## 5. Conclusions

The present study provides preliminary evidence for initial and selective improvements in cortical inhibitory function following 4 weeks of intensive aerobic exercise training in people with chronic stroke. Our results provide a basis for future studies to investigate effects of exercise metabolites such as lactate on post-stroke inhibitory control and associated inhibitory cortical plasticity. Our findings support a multimodal approach that synergizes aerobic exercise and skilled practice targeting complementary cortical inhibitory mechanisms to maximize post-stroke neurorehabilitation outcomes.

## Data Availability

The datasets generated and analyzed supporting the conclusions of this article will be made available by the authors upon request, without undue reservation.

## Acknowledgements

This work was supported by the Eunice Kennedy Shriver National Institute of Child Health & Human Development and the National Institute on Aging of the National Institutes of Health [NIH K99AG075255 (JP), R01HD093694 (PB, DR, SB), P30 AG072973 (JP, SB), P30AG035982 (JP, SB), UL1TR000001, F32MH129076 (AP), T32HD057850 (AW)], the American Heart Association [898190 (AW)], and the Georgia Holland Endowment Fund. The content of this publication is solely the responsibility of the authors and does not necessarily represent the official views of the National Institutes of Health or any other funding agency. We would like to acknowledge Doctor of Physical Therapy students Preston Judd, Katelyn Struckle, and Kailee Carter for their assistance in delivery of the exercise intervention and data analyses.

## Conflict of Interest

*The authors declare that the research was conducted in the absence of any commercial or financial relationships that could be construed as a potential conflict of interest*.

## 4 Supplementary Materials

**Supplemental Figure S1.**
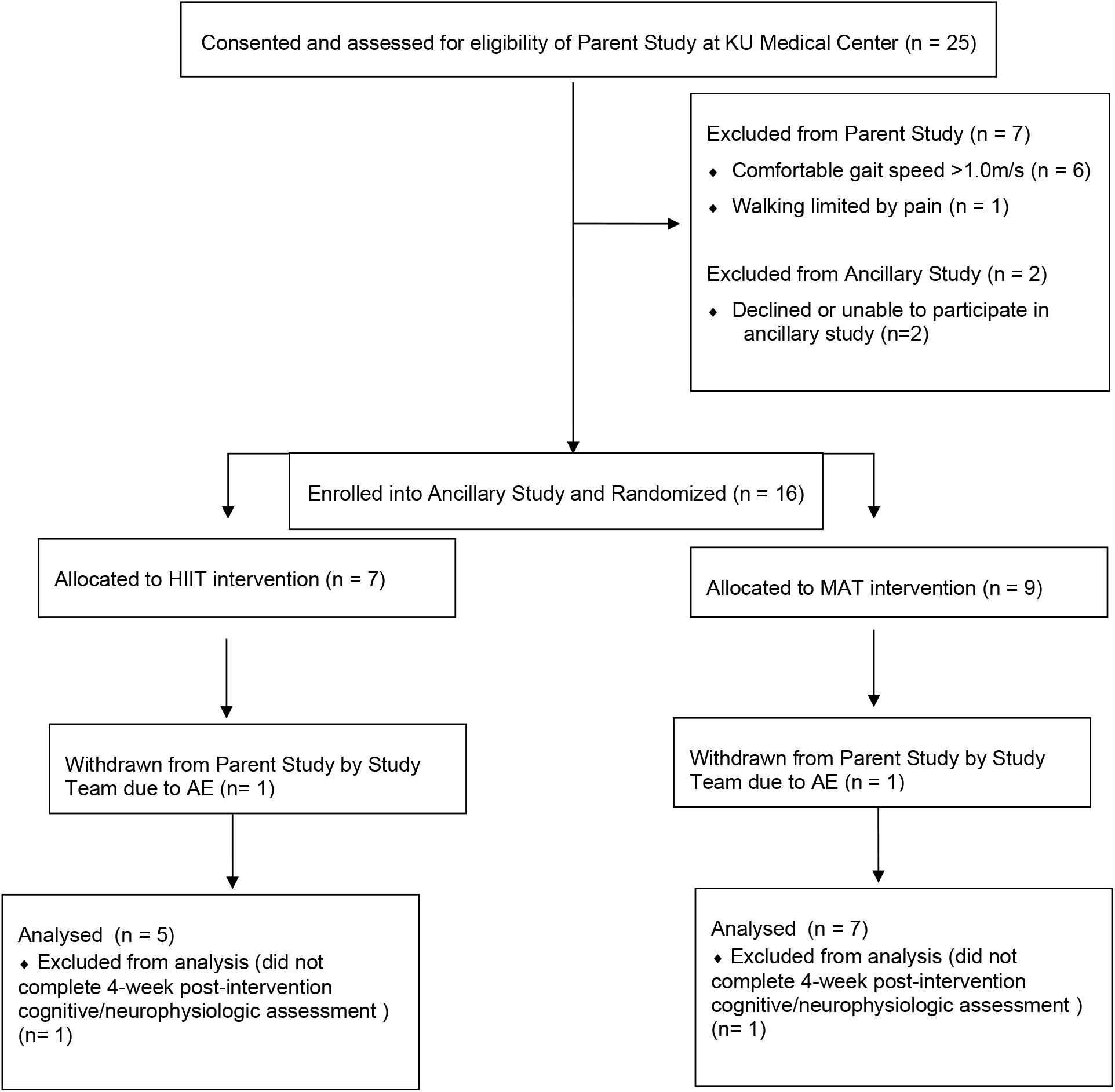
CONSORT Diagram.

## Notes

### Competing Interest Statement

The authors have declared no competing interest.

### Clinical Trial

NCT03760016

### Author Declarations

University of Kansas Institutional Review Board

